# Protection against Omicron BA.2 reinfection conferred by primary Omicron or pre-Omicron infection with and without mRNA vaccination

**DOI:** 10.1101/2022.06.23.22276824

**Authors:** Sara Carazo, Danuta M. Skowronski, Marc Brisson, Sapha Barkati, Chantal Sauvageau, Nicholas Brousseau, Rodica Gilca, Judith Fafard, Denis Talbot, Manale Ouakki, Vladimir Gilca, Alex Carignan, Geneviève Deceuninck, Philippe De Wals, Gaston De Serres

## Abstract

**Background:** We estimated the protection against the Omicron BA.2 variant associated with prior primary infection (PI) due to pre-Omicron or Omicron BA.1 virus, with and without mRNA vaccination.

**Methods:** A test-negative case-control study was conducted among healthcare workers (HCWs) tested for SARS-CoV-2 in Quebec, Canada, between March 27 and June 4, 2022, when BA.2 predominated and was presumptively diagnosed. Logistic regression models compared the likelihood of BA.2 reinfection (second positive test ≥30 days after PI) among HCWs with history of PI and none to three doses of mRNA vaccine versus infection-naïve, unvaccinated HCWs.

**Findings:** Among 37,732 presumed BA.2 cases, 2,521 (6.7%) and 659 (1.7%) were reinfections following pre-Omicron or BA.1 PI, respectively. Among 73,507 controls, 7,360 (10.0%) and 12,315 (16.8%) had a pre-Omicron or BA.1 PI, respectively. Pre-Omicron PI was associated with 38% (95%CI:19-53) reduction in BA.2 infection risk, with higher BA.2 protection among those also vaccinated with one (56%), two (69%) or three (70%) vaccine doses. Omicron BA.1 PI was associated with greater protection against BA.2 (72%; 95%CI:65-78), higher among two-dose vaccinated at 96% (95%CI:95-96) but not improved with a third dose (96%; 95%CI:95-97). Hybrid Omicron BA.1 PI plus two or three dose vaccine-induced protection persisted for five months post-infection.

**Interpretation:** Twice-vaccinated individuals who experienced BA.1 infection were subsequently well-protected for a prolonged period against BA.2 reinfection and derived no meaningful added benefit against BA.2 from a third dose of mRNA vaccine.

## Introduction

Since December 2021, the dominant SARS-CoV-2 variant globally has been Omicron (B.1.1.529), responsible for the highest COVID-19 incidence rates worldwide as a result of its greater transmissibility and escape from natural and vaccine-induced immunity.^1,2^ The initial Omicron BA.1 sublineage has been replaced by the phylogenetically distinct and yet more transmissible BA.2 sublineage.^3–5^ In Quebec, Canada, Omicron BA.2 became dominant at end of March 2022, accounting for >90% of sequenced viruses during the ensuing weeks.^6^

The intense Omicron BA.1 surge that occurred among the highly-vaccinated Quebec population between December 2021 and February 2022,^7^ resulted in a considerable pool of people with potential hybrid immunity induced by the combination of vaccination and infection. In the context of previous reports of reduced and rapidly waning vaccine effectiveness (VE) against Omicron and its sublineages,^1,8–10^ the potential benefit of booster doses requires updated understanding of the protection against Omicron BA.2 conferred by natural and hybrid immunity associated with primary infection (PI) with pre-Omicron or Omicron BA.1 virus.

We used the population-based cohort of all Quebec healthcare workers (HCWs) to estimate protection against Omicron BA.2 reinfection conferred by a pre-Omicron or Omicron BA.1 primary infection, with and without history of mRNA vaccination.

## Methods

### Study design

This test-negative case-control study compared HCWs with a nucleic acid amplification testing (NAAT) positive result for SARS-CoV-2 from March 27 to June 4, 2022 (cases) to HCWs with a negative result during the same period (controls). Tests done within 30 days of a positive result and negative tests collected ≤7 days before a positive specimen (during the incubation period) were excluded. One randomly-selected negative specimen per person contributed as control.

### Population and data sources

The study population included all HCWs paid by the Quebec publicly-funded healthcare system and/or registered as members of a health college (physicians, nurses, nursing assistants, respiratory therapists, midwifes and pharmacists). We excluded participants who were <18 years old, had received four or more vaccine doses before specimen collection, had a first reinfection detected before the specimen collection date, were recipients of a non-mRNA vaccine (AstraZeneca or Janssen), were tested for confirmation of recovery, had an invalid vaccination date, had received a first dose <14 days or a second or third dose <7 days before specimen collection, with the first and second doses administered <21 days apart or the second and third doses administered <90 days apart, as recommended for the general population of Quebec.^11^ We additionally excluded cases identified by epidemiological link or rapid antigen detection testing (RADT) without NAAT confirmation.

We identified cases and controls using the provincial laboratory database that records all NAAT for SARS-CoV-2 in Quebec, including the date of specimen collection, the result and the testing indication. Testing indications include: (1) symptomatic during consultation at the emergency room or hospitalization; (2) other symptomatic HCWs; (3) asymptomatic in the context of outbreaks; (4) hospital pre-admission screening; (5) contact with cases; (6) confirmation of a RADT positive result; (7) confirmation of recovery; and (8) other reasons combined. NAAT has been broadly accessible to HCWs in Quebec throughout the pandemic.

Using a unique personal identifying number, the cohort of HCWs was linked with (1) the laboratory database; (2) the provincial immunization registry, which is a population-based database including all unvaccinated and vaccinated individuals with their dates of vaccination and the type of vaccine administered in Quebec; (3) the provincial database of all COVID-19 cases, including demographic and clinical information; and (4) the administrative hospitalization database.

### Primary infection, vaccination and outcome definitions

We defined exposure by a combination of prior infection and vaccination history. We defined prior primary infection as a NAAT positive specimen collected at least 30 days before a specimen collected during the study period. The 30-day interval was chosen to capture all potential BA.2 reinfections following prior BA.1 PI; however, sensitivity analysis using the more standard 90-day interval to define reinfection was also undertaken.

We defined a pre-Omicron PI as any positive specimen collected between February 20, 2020 and November 27, 2021. The strategy for variant of concern (VOC) identification in Quebec during this period has been detailed elsewhere.^12^ Based on provincial-level genomic surveillance and percentage VOC contribution, we assumed Omicron BA.1 attribution for all cases between December 26, 2021 and March 26, 2022. We excluded participants with a past infection during the period of Delta and Omicron co-circulation (November 28 to December 25, 2021).

We defined vaccination as the administration of BNT162b2 (Pfizer-BioNTech) or mRNA-1273 (Moderna) (mRNA) vaccines or a combination of both. HCWs with pre-Omicron or Omicron BA.1 subvariant primary infection and no vaccine (PI-NV) or one (PI-V1), two (PI-V2) or three (PI-V3) vaccine doses were compared to previously non-infected, non-vaccinated individuals (NI-NV).

The main outcomes were any SARS-CoV-2 infection or symptomatic infections only (identified by specified testing indication) confirmed by NAAT during the study period determined by Omicron BA.2 subvariant dominance when 86% (60% to 93%) of viruses from sentinel laboratories characterized by sequencing were BA.2 (Figure 1).^6^

**Figure 1.**
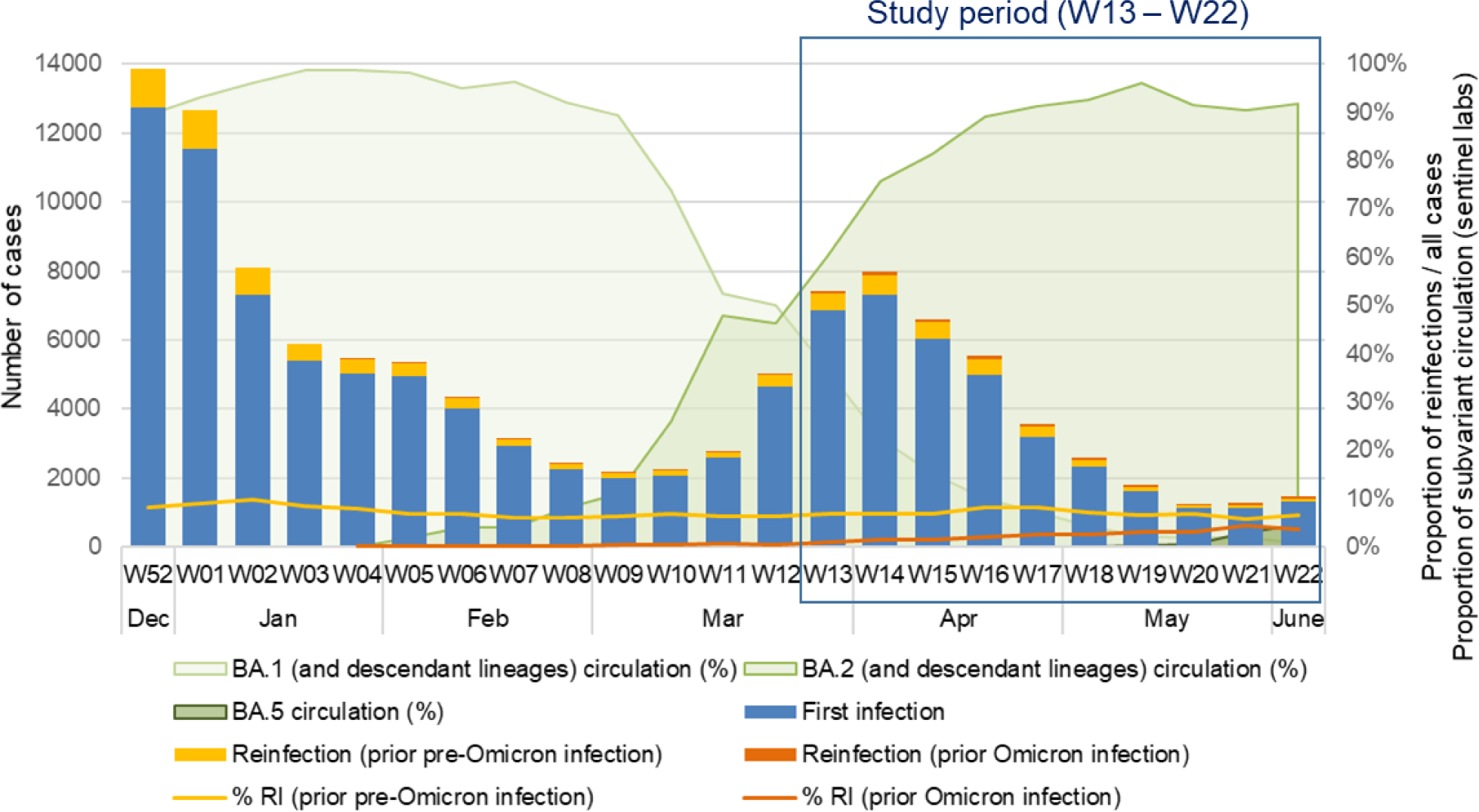
Weekly distribution of SARS-CoV-2 infections by infection history and weekly proportion of reinfections among all infections during the Omicron BA.1 and BA.2 waves. Abbreviations: RI, reinfection; W, epidemiological week

### Statistical analysis

The odds of pre-Omicron or Omicron BA.1 PI with/without vaccination or of vaccination alone without PI were compared among cases and controls. The comparator group for all analyses were NI-NV individuals. Adjusted odds ratios (aOR) and their 95% confidence intervals (CI) were estimated using logistic regression models adjusted for age, sex, type of employment, facility, testing indication and epidemiological week. Effectiveness/protection was derived as 1 – aOR. ORs were not estimated for exposure categories with less than five individuals. Analyses were also stratified by time since last PI or vaccine exposure.

Analyses were performed using SAS version 9.4 (SAS Institute Inc).

### Ethic statement

This study was conducted under the legal mandate of the National Director of Public Health of Quebec under the Public Health Act, granting a participant consent waiver. It was also approved by the Research Ethics Board of the Centre hospitalier universitaire de Québec-Université Laval.

## Results

### Study population

Among the 258,007 tests performed among HCWs during the study period, 99.5% were merged with other databases. Reasons for exclusion of 15.5% of the tests are detailed in Supplementary Figure 1. In total, 37,732 presumptive Omicron BA.2 cases were compared to 73,507 randomly-selected controls. Tables 1 and 2 describe the characteristics and vaccination status of cases and controls by PI history. Most of participants were adults aged 18 to 59 years old (94.1%) and women (82.6%). A prior SARS-CoV-2 PI was detected in 8.4% and 26.8% of cases and controls, respectively (Table 2). PI was combined with two or three vaccine doses in 3.1% and 3.8% of cases, respectively, and 10.9% and 13.4% of controls, respectively.

**Table 1.** Characteristics of cases and controls stratified by SARS-CoV-2 primary infection (PI) history.

|  | **Cases (n=37732)** |  | **Controls (n=73507)** |  |
| --- | --- | --- | --- | --- |
|  | **Prior primary infection** | **No primary infection** | **Prior primary infection** | **No primary infection** |
| **Total N** | 3180 | 34552 | 19675 | 53832 |
|  | **n (%)** | **n (%)** | **n (%)** | **n (%)** |
| **Sex** |  |  |  |  |
| Female | 2643 (82.8) | 28235 (81.7) | 16566 (84.2) | 44401 (82.5) |
| Male | 546 (17.2) | 6317 (18.3) | 3109 (15.8) | 9431 (17.5) |
| **Age (years)** |  |  |  |  |
| 18-39 | 1750 (55.0) | 16473 (47.7) | 10833 (55.1) | 24383 (45.3) |
| 40-59 | 1341 (42.2) | 16161 (46.8) | 8106 (41.2) | 25629 (47.6) |
| ≥60 | 89 (2.8) | 1918 (5.6) | 736 (3.7) | 3820 (7.1) |
| **Type of employment** |  |  |  |  |
| Physician | 104 (3.3) | 1653 (4.8) | 465 (2.4) | 3014 (5.6) |
| Nursing staff and respiratory therapists | 1220 (38.4) | 10736 (31.1) | 7706 (39.2) | 18051 (33.5) |
| Other health assisting occupations | 980 (30.8) | 8319 (24.1) | 6048 (30.7) | 13857 (25.7) |
| Social workers | 569 (17.9) | 8317 (24.1) | 3464 (17.6) | 11131 (20.7) |
| Pharmacist | 24 (0.8) | 505 (1.5) | 160 (0.8) | 974 (1.8) |
| Management and administrative staff | 283 (8.9) | 5022 (14.5) | 1832 (9.3) | 6805 (12.6) |
| **Facility** |  |  |  |  |
| Hospital / health centers | 1712 (53.8) | 18798 (54.4) | 10927 (55.5) | 29557 (54.9) |
| Long-term health facilities | 605 (19.0) | 3705 (10.7) | 3736 (19.0) | 7020 (13.0) |
| Rehabilitation centers | 141 (4.4) | 1689 (4.9) | 712 (3.6) | 2248 (4.2) |
| Childhood and youth centers | 89 (2.8) | 1547 (4.5) | 561 (2.9) | 1569 (2.9) |
| Home care | 182 (5.7) | 2057 (6.0) | 1023 (5.2) | 2640 (4.9) |
| Other | 451 (14.2) | 6756 (19.6) | 2716 (13.8) | 10798 (20.1) |
| **Year-epi-week of specimen collection (calendar start/end date)** |  |  |  |  |
| 2022-13 to 2022-14 (Mar 27 – Apr 9) | 1102 (34.7) | 13789 (39.9) | 4737 (24.1) | 19310 (35.9) |
| 2022-15 to 2022-16 (Apr 10 – 23) | 1001 (31.5) | 10676 (30.9) | 5141 (26.1) | 14515 (27.0) |
| 2022-17 to 2022-18 (Apr 24 – May 7) | 563 (17.7) | 5284 (15.3) | 4271 (21.7) | 8995 (16.7) |
| 2022-19 to 2022-20 (May 8 – 21) | 264 (8.3) | 2556 (7.4) | 2862 (14.6) | 5983 (11.1) |
| 2022-21 to 2022-22 (May 22 – June 4) | 250 (7.9) | 2247 (6.5) | 2664 (13.5) | 5029 (9.3) |
| **Indication for testing** |  |  |  |  |
| Symptomatic, emergency room | 52 (1.6) | 445 (1.3) | 820 (4.2) | 1636 (3.0) |
| Symptomatic, healthcare worker | 1765 (55.5) | 21058 (60.9) | 7374 (37.5) | 20588 (38.2) |
| Asymptomatic, closed setting outbreak | 252 (7.9) | 1523 (4.4) | 4617 (23.5) | 10767 (20.0) |
| Asymptomatic, hospital pre-admission | 41 (1.3) | 318 (0.9) | 1417 (7.2) | 3880 (7.2) |
| Asymptomatic contact | 283 (8.9) | 3710 (10.7) | 1955 (9.9) | 8749 (16.3) |
| Asymptomatic other | 177 (5.6) | 915 (2.6) | 1973 (10.0) | 4758 (8.8) |
| Confirmation of positive rapid antigenic test | 562 (17.7) | 6324 (18.3) | 938 (4.8) | 2277 (4.2) |
| Other reasons combined | 48 (1.5) | 259 (0.7) | 581 (3.0) | 1177 (2.2) |
| **VOC of PI by calendar period** |  |  |  |  |
| Pre-VOC (before Feb 1, 2021) | 2081 (65.4) | NA | 5996 (30.5) | NA |
| Pre-VOC / Alpha (Feb 1 – Apr 10, 2021) | 185 (5.8) | NA | 503 (2.6) | NA |
| Alpha (Apr 11 – June 26, 2021) | 98 (3.1) | NA | 280 (1.4) | NA |
| Alpha / Delta (June 27 – Sep 4, 2021) | 53 (1.7) | NA | 141 (0.7) | NA |
| Delta (Sep 5 – Nov 27, 2021) | 104 (3.3) | NA | 440 (2.2) | NA |
| Omicron BA.1 (Dec 26, 2021 – Mar 26, 2022) | 630 (19.8) | NA | 11585 (58.9) | NA |
| Omicron BA.2 (Mar 27 – May 7, 2022) | 29 (0.9) | NA | 730 (3.7) | NA |
| **Days between PI and specimen collection**<br>(median, minimum – maximum) | 487 (30 – 801) | NA | 120 (30 – 804) | NA |

Abbreviations: NA, non-applicable; NI, no infection previously; PI, prior primary infection; VOC, variant of concern

**Table 2.** Vaccination status of presumptive Omicron BA.2 cases and controls stratified by SARS-CoV-2 primary infection (pre-Omicron or Omicron BA.1)

|  | All infections analysis |  | Symptomatic infections analysis<sup>a</sup> |  |
| --- | --- | --- | --- | --- |
|  | Cases | Controls | Cases | Controls |
|  | N (%) | N (%) | N (%) | N (%) |
| **Overall** | **37732** | **73507** | **23371** | **30542** |
| **Pre-Omicron PI** | **2521 (6.7)** | **7360 (10.0)** | **1495 (6.4)** | **2643 (8.7)** |
| Non-vaccinated (PI-NV) | 109 (0.3) | 265 (0.4) | 57 (0.2) | 41 (0.1) |
| One vaccine dose | **342 (0.9)** | **729 (1.0)** | **214 (0.9)** | **285 (0.9)** |
| PI-V1 | 340 (0.9) | 722 (1.0) | 212 (0.9) | 283 (0.9) |
| V1-PI | 2 (0.0) | 7 (0.0) | 2 (0.0) | 2 (0.0) |
| Two vaccine doses | **897 (2.4)** | **2665 (3.6)** | **540 (2.3)** | **987 (3.2)** |
| PI-V2 | 815 (2.2) | 2376 (3.2) | 486 (2.1) | 860 (2.8) |
| V1-PI-V2 | 39 (0.1) | 107 (0.1) | 24 (0.1) | 56 (0.2) |
| V2-PI | 43 (0.1) | 182 (0.2) | 30 (0.1) | 71 (0.2) |
| Three vaccine doses | **1173 (3.1)** | **3701 (5.0)** | **684 (2.9)** | **1330 (4.4)** |
| PI-V3 | 1028 (2.7) | 3177 (4.3) | 594 (2.5) | 1118 (3.7) |
| V1-PI-V3 | 83 (0.2) | 271 (0.4) | 52 (0.2) | 99 (0.3) |
| V2-PI-V3 | 62 (0.2) | 253 (0.3) | 38 (0.2) | 113 (0.4) |
| **Omicron BA.1 PI** | **659 (1.7)** | **12315 (16.8)** | **330 (1.4)** | **5600 (18.3)** |
| Non-vaccinated (PI-NV) | 125 (0.3) | 727 (1.0) | 43 (0.2) | 119 (0.4) |
| One vaccine dose | **9 (0.0)** | **109 (0.1)** | **3 (0.0)** | **33 (0.1)** |
| PI-V1 | 1 (0.0) | 6 (0.0) | 1 (0.0) | 2 (0.0) |
| V1-PI | 8 (0.0) | 103 (0.1) | 2 (0.0) | 31 (0.1) |
| Two vaccine doses | **262 (0.7)** | **5322 (7.2)** | **147 (0.6)** | **2519 (8.2)** |
| V2-PI | 262 (0.7) | 5314 (7.2) | 147 (0.6) | 2517 (8.2) |
| V1-PI-V2 | 0 (0.0) | 8 (0.0) | 0 (0.0) | 2 (0.0) |
| Three vaccine doses | **263 (0.7)** | **6157 (8.4)** | **137 (0.6)** | **2929 (9.6)** |
| V3-PI | 243 (0.6) | 5679 (7.7) | 124 (0.5) | 2716 (8.9) |
| V2-PI-V3 | 20 (0.1) | 478 (0.7) | 13 (0.1) | 213 (0.7) |
| **Not infected (NI) previously with SARS-CoV-2** | **34552 (91.6)** | **53832 (73.2)** | **21546 (92.2)** | **22299 (73.0)** |
| Non-vaccinated (NI-NV) | 672 (1.8) | 1043 (1.4) | 343 (1.5) | 125 (0.4) |
| One vaccine dose (NI-V1) | 136 (0.4) | 193 (0.3) | 68 (0.3) | 45 (0.1) |
| Two vaccine doses (NI-V2) | 6717 (17.8) | 8939 (12.2) | 4387 (18.8) | 3839 (12.6) |
| Three vaccine doses (NI-V3) | 27027 (71.6) | 43657 (59.4) | 16748 (71.7) | 18290 (59.9) |

Abbreviations: NI, not infected previously; NI-NV, not infected previously non-vaccinated; NI-V1, not infected previously, one vaccine dose; NI-V2, not infected previously, two vaccine doses; NI-V3, not infected previously, three vaccine doses; PI, prior primary infection; PI-NV, prior infection non-vaccinated; PI-V1, prior infection before one vaccine dose; PI-V2, prior infection before two vaccine doses; PI-V3, prior infection before three vaccine doses; V1-PI, prior infection after one vaccine dose; V1-PI-V2, prior infection after first but before second vaccine dose; V1-PI-V3, prior infection after first but before second and third vaccine doses; V2-PI, prior infection after two vaccine doses; V2-PI-V3, prior infection after second but before third vaccine dose; V3-PI, prior infection after three vaccine doses
<sup>a</sup>Cases symptomatic at the moment of specimen collection according to the indication for testing

COVID-19 hospitalizations within 30 days after specimen collection were recorded for 59 vaccinated cases: 51 among cases without PI (0.2%), four among cases with pre-Omicron PI (0.2%) and three among cases with BA.1 PI (0.6%). Only one COVID-19-attributable death was documented in a three-dose vaccinated non-previously infected individual (data not shown).

### Omicron BA.2 reinfections

Among the 2,521 (6.7%) cases reinfected after pre-Omicron PI, most were vaccinated following their PI (32.3% PI-V2 and 40.8% PI-V3 respectively) and 4.3% remaining unvaccinated. Among the 659 (1.7%) cases reinfected after a presumptive Omicron BA.1 PI, 19.0% were nonvaccinated, and 39.8% and 36.9% had been vaccinated before the PI with two and three doses, respectively (Table 2).

Reinfections with a prior pre-Omicron PI mostly (82.3%) occurred before February 2021 during the pre-VOC period, with a median PI to reinfection interval of 515 days (range: 127-801) (Figure 2). Reinfections with a prior BA.1 PI, mostly (65%) occurred during the three peak weeks of the first Omicron BA.1 wave, with a median PI to reinfection interval of 100 days (range: 30-158). Among them, 49 (6.5%) occurred at 30-59 days post-PI and 151 (20.8%) at 60-89 days post-PI (figure 2). The proportion of reinfections following a BA.1 PI increased from 0.9% to 3.6% of the reported cases from epi-weeks 13 to 22 of 2022 (Figure 1).

**Figure 2.**
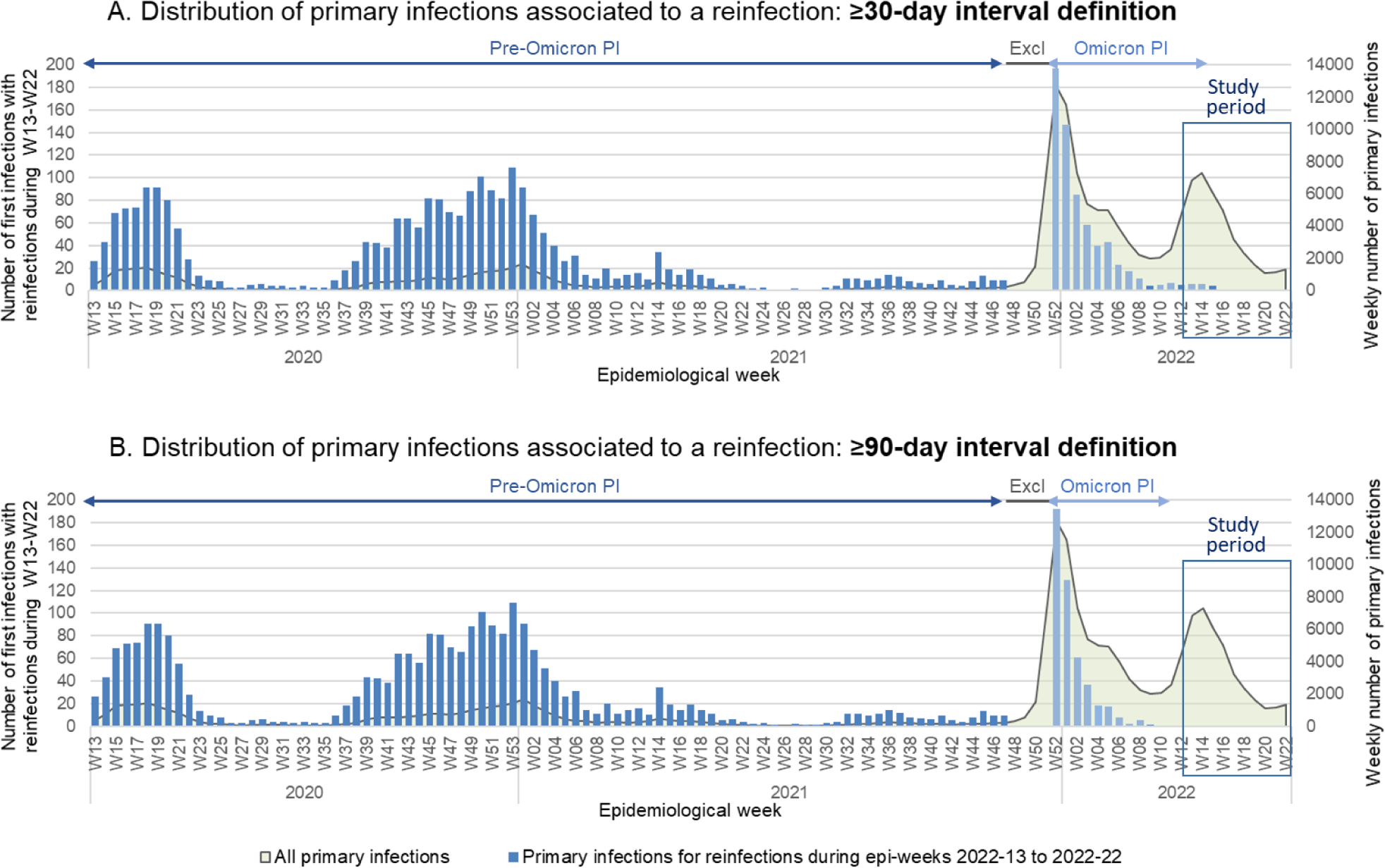
Distribution of primary infections relative to reinfections during the study period, by definition of reinfection (≥30-day versus ≥90-day interval) Abbreviations: PI, prior primary infection; Excl, excluded period; W, epidemiological week

### Pre-Omicron PI-induced protection with/without vaccination

Pre-Omicron PI alone (without vaccination) was associated with a reduced risk of any BA.2 reinfection of 38% (95%CI: 19-53) and of symptomatic reinfection of 51% (95%CI: 22-69). In those with two-dose plus pre-Omicron PI hybrid exposure, the estimated protection was 69% (95%CI: 64-73), similar to that with three-dose plus PI exposure (70%, 95%CI: 66-74). Estimated pre-Omicron hybrid protection against symptomatic BA.2 reinfection was also similar with two (81%, 95%CI: 76-85) or three vaccine doses (83%, 95%CI: 78-86) (Figure 3 and Supplementary Table 1).

**Figure 3.**
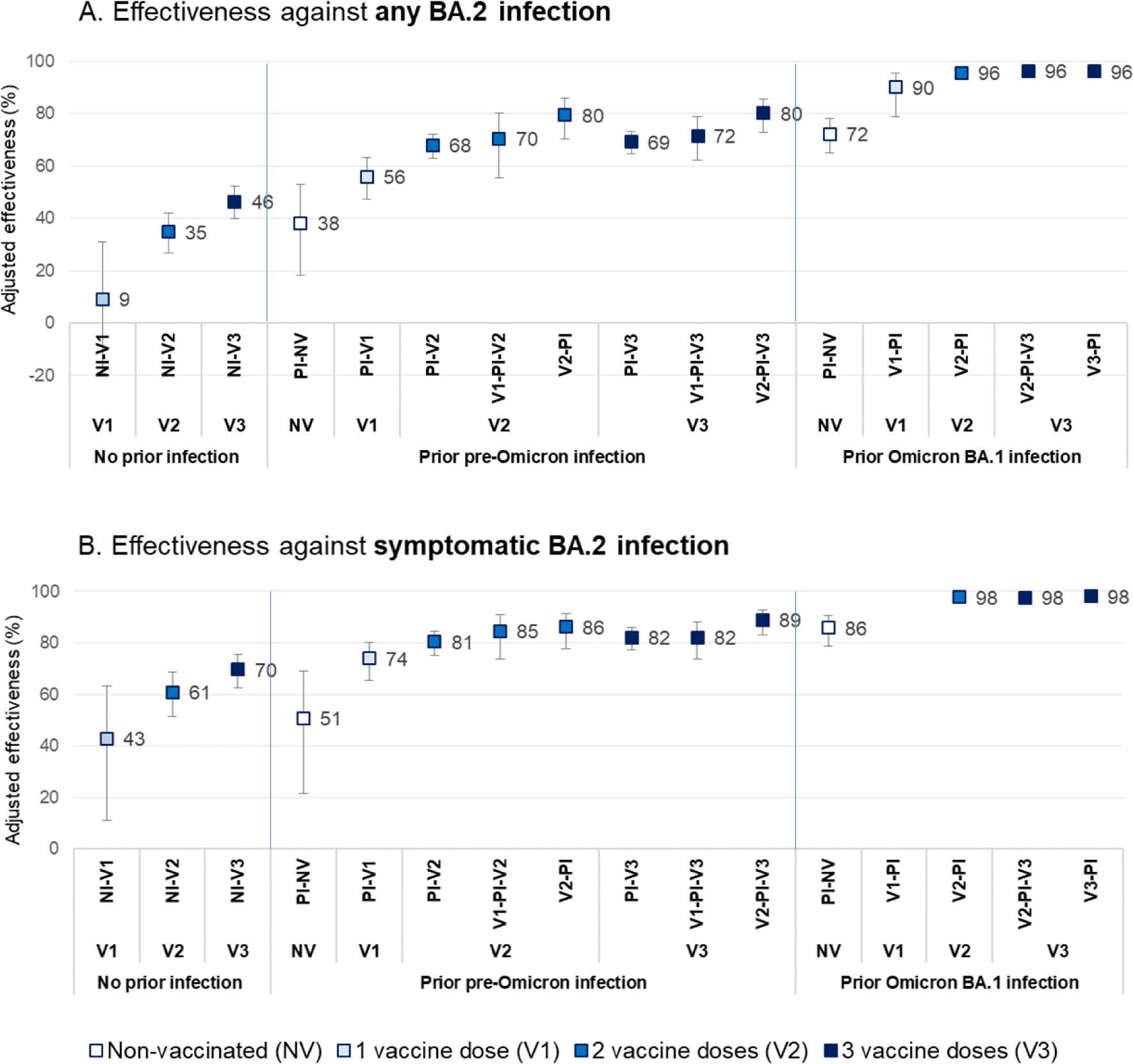
Effectiveness against Omicron BA.2 infection (any infection or symptomatic infection) conferred by pre-Omicron or Omicron BA.1 primary infection with/without vaccination. Abbreviations: NI, not infected previously; NI-V1, not infected previously, one vaccine dose; NI-V2, not infected previously, two vaccine doses; NI-V3, not infected previously, three vaccine doses; PI, prior primary infection; PI-NV, prior infection non-vaccinated; PI-V1, prior infection before one vaccine dose; PI-V2, prior infection before two vaccine doses; PI-V3, prior infection before three vaccine doses; V1-PI, prior infection after one vaccine dose; V1-PI-V2, prior infection after first but before second vaccine dose; V1-PI-V3, prior infection after first but before second and third vaccine doses; V2-PI, prior infection after two vaccine doses; V2-PI-V3, prior infection after second but before third vaccine dose; V3-PI, prior infection after three vaccine doses Note: Logistic regression models comparing persons with prior primary infection and/or vaccination to those unvaccinated and without prior infection. All estimates adjusted for age, sex, type of employment, facility, indication for testing and epidemiological week

### Omicron BA.1 PI-induced protection with/without vaccination

Omicron BA.1 PI alone (without vaccination) was associated with a risk reduction of any BA.2 reinfection of 72% (95%CI: 65-78) and of symptomatic reinfection of 86% (95%CI: 79-91). This protection was similar to that of hybrid pre-Omicron PI plus two or three vaccine doses and higher than the estimated three-dose vaccine effectiveness (46%; 95%CI: 40-52) among the previously non-infected (Figure 3). This difference was not explained by time since last exposure. At similar 3 to <6-month interval since PI, Omicron BA.1 PI without vaccination was associated with a BA.2 risk reduction of 72% compared to 32% for pre-Omicron PI without vaccination (Table 3). At similar 30 to 89-days interval since last exposure, Omicron BA.1 PI without vaccination was associated with BA.2 risk reduction of 78% (95%CI: 66-86) compared to 63% (95%CI: 57-68) for three vaccine doses without prior infection (data not shown).

**Table 3.** Protection against any Omicron BA.2 reinfection associated with pre-Omicron or Omicron BA.1 primary infection with/without vaccination by time since last immunogenic event (vaccination or primary infection)

|  | Infection history |  |  |  |
| --- | --- | --- | --- | --- |
|  | Pre-Omicron PI |  | Omicron BA.1 PI |  |
| Time since last immunogenic event before testing: | Unadjusted effectiveness<sup>a</sup><br>(95% CI) | Adjusted effectiveness<sup>a,b</sup><br>(95% CI) | Unadjusted effectiveness<sup>a</sup><br>(95% CI) | Adjusted effectiveness<sup>a,b</sup><br>(95% CI) |
| **Time since PI among unvaccinated (PI-NV)** |  |  |  |  |
| 30 – 59 days (1 –<2 months) | NE | NE | 78% (43, 91) | 82% (49, 94) |
| 60 – 89 days (2 –<3 months) | NE | NE | 72% (59, 82) | 76% (63, 85) |
| 90 – 182 days (3 –<6 months) | 13% (-99, 62) | 42% (-47, 77) | 73% (66, 79) | 70% (61, 77) |
| 183 – 364 days (6 –<12 months) | 38% (5, 60) | 39% (0, 63) | NE | NE |
| 365 - 757 days (≥12 months) | 37% (16, 53) | 42% (17, 60) | NE | NE |
| **Time since PI among 2-dose vaccinated (V2-PI)** |  |  |  |  |
| 30 – 59 days (1 –<2 months) | NE | NE | 94% (88, 97) | 97% (94, 98) |
| 60 – 89 days (2 –<3 months) | NE | NE | 93% (90, 95) | 97% (96, 98) |
| 90 – 159 days (3 –<6 months) | NE | NE | 92% (91, 94) | 96% (95, 96) |
| **Time since PI among 3-dose vaccinated (V3-PI)** |  |  |  |  |
| 30 – 59 days (1 –<2 months) | NE | NE | 93% (89, 95) | 96% (94, 98) |
| 60 – 89 days (2 –<3 months) | NE | NE | 93% (91, 95) | 97% (96, 98) |
| 90 – 158 days (3 –<6 months) | NE | NE | 94% (92, 95) | 96% (95, 97) |
| **Time since second vaccine dose (PI-V2 or V1-PI-V2)** |  |  |  |  |
| 7 – 59 days (<2 months) | 71% (48, 84) | 89% (78, 94) | NE | NE |
| 60 – 89 days (2 –<3 months) | 42% (18, 59) | 73% (60, 82) | NE | NE |
| 90 – 182 days (3 –<6 months) | 59% (50, 66) | 77% (71, 82) | NE | NE |
| 183 – 364 days (6 –<12 months) | 41% (32, 48) | 68% (62, 74) | NE | NE |
| **Time since third vaccine dose (PI-V3 or V1-PI-V3 or V2-PI-V3)** |  |  |  |  |
| 7 – 59 days (<2 months) | 71% (55, 80) | 88% (81, 92) | 94% (90, 97) | 98% (96, 99) |
| 60 – 89 days (2 –<3 months) | 49% (39, 57) | 80% (75, 84) | 90% (78, 96) | 95% (89, 98) |
| 90 – 182 days (3 –<6 months) | 50% (44, 56) | 72% (67, 76) | NE | NE |
| 183 – 305 days (6 –<10 months) | 74% (-115, 97) | 82% (-109, 98) | NE | NE |

<sup>a</sup> Logistic regression models comparing persons with prior primary infection with/without vaccination to those unvaccinated and without prior infection, relative to unvaccinated individuals with no infection history
<sup>b</sup> Estimates adjusted for age, sex, type of employment, facility, indication for testing and epidemiological week.
Abbreviations: CI, confidence interval; NE, non-estimable; PI, prior primary infection; PI-NV, prior infection non-vaccinated; PI-V1, prior infection before one vaccine dose; PI-V2, prior infection before two vaccine doses; PI-V3, prior infection before three vaccine doses; V1-PI-V2, prior infection after first but before second vaccine dose; V1-PI-V3, prior infection after first but before second and third vaccine doses;; V2-PI-V3, prior infection after second but before third vaccine dose

Omicron BA.1 PI plus two-dose hybrid exposure was associated with 96% (95%CI: 95-96) and 98% (95%CI: 97-98) reduced risk of any or symptomatic Omicron BA.2 reinfection, respectively. As observed with pre-Omicron PI, the estimated hybrid BA.1 PI protection was not improved with a third vaccine dose against any (96%, 95%CI: 95-97) or symptomatic (98%, 95%CI: 98-99) infection (Supplementary Table 1). The timing of the third dose prior (V3-PI) or following (V2-PI-V3) the PI did not modify the observed protection (Figure 3). No differences were found in sensitivity analyses using the more standard 90-day interval for defining reinfection (Supplementary Table 2).

### Duration of protection

Pre-Omicron PI without vaccination, occurring four to 25 months before Omicron BA.2 circulation, was associated with 42%, 39% and 42% protection against reinfections four to five, six to 11 and ≥12 months later, with wide and overlapping CIs (Table 3). When the last vaccine dose followed the pre-Omicron PI, two and three doses were associated with similar protection during the seven to 59-day post-vaccination period (89% and 88% respectively) or the 60 to 89-day follow-up (73% and 80% respectively).

Over the five months of follow-up among participants with history of prior Omicron BA.1 PI, a non-significant decline in protection was observed among unvaccinated individuals (from 82% at 30-59 days to 70% at 90-160 days), but not among those also vaccinated with two or three doses before their PI, whose protection remained between 96% and 97% for the 30-159 days of follow-up (Table 3).

Estimated protection against reinfection when the third dose was instead administered after an Omicron BA.1 PI was comparably high at 98% during the 7-59 days and 95% during the 60-89 days following their third dose. No two-dose vaccinated individuals had received their second dose after Omicron BA.1 PI (Table 3).

## Discussion

Prior Omicron BA.1 primary infection alone was the single-most protective event against any (72%) or symptomatic (86%) BA.2 infection, higher than conferred by a pre-Omicron primary infection alone (38% and 51%, respectively) or by even three doses of mRNA vaccine in infection-naïve individuals (46% and 70%, respectively). Hybrid immunity conferred by prior Omicron BA.1 infection plus two or three vaccine doses similarly increased the estimated effectiveness to 96%. Protection was maintained for at least five months post-infection. When the priming infection was a pre-Omicron SARS-CoV-2, the risk reduction of BA.2 reinfection was 38% in unvaccinated individuals, but >85% in those who had received their second or third vaccine dose less than two months earlier, and >70% when these doses were administered two to six months earlier.

In our population of HCWs, 1.7% of cases detected during the BA.2-dominant study period had been previously infected by the Omicron BA.1 variant, a risk four times lower than among those previously infected by pre-Omicron SARS-CoV-2 virus, but still higher than reported reinfection rates before the Omicron surge.^13–15^ The Omicron BA.2 sublineage shares multiple mutations with BA.1, but with genetic differences conferring growth advantage and resulting in BA.1 displacement.^3^ Omicron BA.2 reinfections had been documented using whole genome sequencing as early as 20 days following an Omicron BA.1 primary infection, although such occurrences appeared rare.^16,17^

We observed a moderate protection against any (38%) and symptomatic infection (51%) by heterologous pre-Omicron primary infection without vaccination, but a higher estimate for two-dose hybrid immunity (81%). An study from Qatar reported a 46% effectiveness against symptomatic BA.2 infection in individuals with pre-Omicron PI and 55% and 77% if combined with two- or three-dose vaccination, respectively.^18^ Protection was >70% against any severe outcome. Their lower estimate for two dose hybrid immunity compared to our results might be potentially explained by a longer time since second dose. Andeweg et al reported in a preprint that effectiveness against BA.2 infection for individuals previously (pre-Omicron or Omicron) infected was 60-80% for two-dose vaccinated less than one to seven months earlier, and 75-80% for three-dose vaccinated less than one to four months earlier.^19^ Studies examining protection against Omicron reinfection (BA.1 or any sublineage) have reported that prior pre-Omicron on infection was associated with moderate effectiveness (25% to 47%), but stronger against severe outcome (>80%), and improved with vaccination.^12,18–22^ As reported in our study, a third dose among previously infected individuals was associated to a transient increase in protection both against Omicron BA.1 and BA.2 subvariants.^12,19,20^

Evidence about cross-protection between different Omicron sublineages is scarce. Preliminary immunologic data show robust neutralizing antibody titers against BA.2 in sera of previously infected by BA.1, that improved in those also vaccinated.^23,24^ Few studies to date have estimated real-word cross-protection. We found that prior Omicron infection alone was associated with 72% protection against BA.2 reinfection, and 82% when primary infection occurred 30 to 59 days earlier. In a preprint, Chemaitelly et al evaluated cross-sublineage protection against BA.2 reinfection ≥35 days later and reported that BA.1 was 95% protective against BA.2 reinfection during one to 60 days of follow up.^25^ Authors adjusted for but did not stratify by vaccine status, failing to elucidate the difference in protection with natural versus hybrid immunity. In their preprint, Andeweg et al reported an estimated ∼70% effectiveness against BA.2 reinfection when the primary infection (without vaccination) was 30 to 59 days earlier, during Omicron-dominant circulation and thus indirectly measuring BA.1 – BA.2 cross-sublineage protection.^19^ Authors did not, however, report on two- or three-dose hybrid protection for primary infections occurring during the Omicron period.

In Quebec, as in many other countries with high vaccine coverage, most cases during the Omicron BA.1 surge were two-dose recipients as booster campaigns for the general population unfolded only in response to signals of Omicron immune evasion and surge.^7^ This created a pool of recently infected and/or recently vaccinated individuals with hybrid immunity that was more closely homologous for the infection-induced component. In our population of HCWs, prior Omicron infection combined with two or three vaccine doses was associated with 96% protection against BA.2. Estimated protection was slightly higher against symptomatic infections, and only three hospitalizations were recorded. No other studies to date have directly examined the effect and duration of protection from hybrid BA.1 immunity against descendant Omicron sublineages.

Reported protection against Omicron reinfection conferred by prior pre-Omicron infection waned with time since the last immunogenic event (primary infection or vaccination).^12,20,26^ However, data interpretation is challenging due to overlapping changes in SARS-CoV-2 variant circulation, vaccination deployment and time since the priming event (primary infection or vaccination). In our study, we observed waning in the first six months from receipt of the second or third vaccine dose among those with heterologous prior pre-Omicron primary infection (from 88-89% to 72-77%), but not for individuals primed with Omicron BA.1 infection during the shorter five-month follow-up period (stable from 96% to 97%). To our knowledge, no studies with longer follow-up have examined the duration of protection from natural or hybrid immunity conferred by Omicron BA.1 primary infection.

Our study has some limitations. We assigned BA.1 and BA.2 sublineages based on calendar time and provincial phylogenetic analysis and surveillance.^6^ We excluded weeks of Delta-Omicron cocirculation to specifically measure the effect of prior Omicron infection, but still ∼14% of infections during the study period might have been due to the Omicron BA.1 sublineage,^6^ which may overestimate Omicron PI protection against BA.2 reinfection. We defined reinfections based on a ≥30-day interval between positive tests because of data documenting early reinfections during SARS-CoV-2 variant replacement periods.^16,17^ Prolonged viral shedding might have been misclassified as reinfections,^27^ but this would tend to underestimate the protection associated with prior BA.1 infection. Reassuringly, sensitivity analysis using the standard ≥90-day interval did not change our estimates of protection. Asymptomatic or pauci-symptomatic infections may have been undetected before or during Omicron waves, which would also lead to underestimation of the protection induced by past infection. Infection ascertainment bias should be low among the prioritized and repeatedly tested HCWs who had easy and continuous access to NAAT testing. Due to the low COVID-19 hospitalization rate among HCWs, the effect of PI severity as well as effectiveness against severe outcomes could not be estimated. Previous data indicate that protection should be higher and longer lasting against severe Omicron disease.^12,20^ The proportion of infection-naïve unvaccinated HCWs, the comparator group for all analyses, was low (<2%), and unmeasured characteristics or exposure behaviours could differ from the other groups leading to residual confusion bias. Reassuringly, our estimates of vaccine and pre-Omicron PI effectiveness are in line with published evidence. Finally, the study was conducted among an actively-working adult population such that results may not be generalizable to children, elderly or immune-compromised adults. Extrapolation of our results to newly circulating Omicron BA.4 and BA.5 or other sublineages requires caution. BA.1 infection-induced neutralizing immunity seems less protective against newly circulating Omicron BA.4 and BA.5 sublineages than BA.2 sublineage, but will likely protect against symptomatic or severe infection with these newly evolving Omicron sublineages.^28,29^ Ongoing evaluation of heterologous cross-protection (infection and hybrid infection plus vaccine induced) against emerging dominant variants and subvariants remains important in informing real-time vaccine program adjustments.

In the context of billions of people worldwide already infected with SARS-CoV-2, over half having accrued since Omicron emergence alone, and with much of the global population being now also twice vaccinated,^30^ our results have important implications for preparedness for potential future waves and timely informing immunization programs. If our findings of substantial and sustained Omicron BA.1 hybrid protection against BA.2 also apply to other emerging Omicron subvariants, then booster doses for twice-vaccinated individuals already infected by BA.1 or BA.2 are likely to be of marginal benefit, particularly against severe outcomes. Such doses may be better oriented toward protecting the more vulnerable globally.

## Conclusion

Previous Omicron BA.1 infection was the single-most protective immunogenic event against Omicron BA.2 infection, with protection against BA.2 enhanced to 96% in those who also received two doses of mRNA vaccine.

## Data Availability

The databases used in this study are a property of the Mininstry of Health of Quebec and were shared with the researchers under the legal mandate of the National Director of Public Health of Quebec under the Public Health Act, preventing data sharing with a third party. Aggregate data are available within the manuscript and the supplementary material.

## Acknowledgments

This work was supported by the Ministère de la santé et des services sociaux du Québec. The authors thank Radhouene Doggi, Abakar Idriss-Hassan and Hany Geagea (Institut national de santé public du Québec) for their contribution in the literature search.

