## Supplemental material for "Protection against Omicron BA.2 reinfection conferred by primary Omicron or pre-Omicron infection with and without mRNA vaccination"

**Supplementary Table 1. Protection against any Omicron BA.2 reinfection and against symptomatic Omicron BA.2 reinfection associated with pre-Omicron and Omicron BA.1 primary infection (PI) with/without vaccination.**

| | Any Omicron BA.2 infection | | Symptomatic Omicron BA.2 infection | |
|---|---|---|---|---|
| **Infection history and vaccination status[a]** | Unadjusted protection (95% CI) | Adjusted protection[b] (95% CI) | Unadjusted protection (95% CI) | Adjusted protection[b] (95% CI) |
| **Pre-Omicron PI** | | | | |
| Unvaccinated | 36% (19, 50) | 38% (19, 53) | 49% (21, 68) | 51% (22, 69) |
| 1-dose vaccinated | 27% (15, 38) | 56% (47, 63) | 73% (64, 79) | 74% (66, 80) |
| 2-dose vaccinated | 48% (41, 54) | 69% (64, 73) | 80% (75, 84) | 81% (76, 85) |
| 3-dose vaccinated | 51% (45, 56) | 70% (66, 74) | 81% (77, 85) | 83% (78, 86) |
| **Omicron BA.1 PI** | | | | |
| Unvaccinated | 73% (67, 78) | 72% (65, 78) | 87% (80, 91) | 86% (79, 91) |
| 1-dose vaccinated | 87% (75, 94) | 89% (78, 95) | 97% (89, 99) | 97% (89, 99) |
| 2-dose vaccinated | 92% (91, 94) | 96% (95, 96) | 98% (97, 98) | 98% (97, 98) |
| 3-dose vaccinated | 93% (92, 94) | 96% (95, 97) | 98% (98, 99) | 98% (98, 99) |
| **No prior infection** | | | | |
| Unvaccinated | Referent | Referent | Referent | Referent |
| 1-dose vaccinated | -9% (-39, 14) | 9% (-19, 31) | 45% (16, 64) | 43% (11, 63) |
| 2-dose vaccinated | -17% (-29, -5) | 35% (27, 42) | 58% (49, 66) | 61% (52, 69) |
| 3-dose vaccinated | 4% (-6, 13) | 46% (40, 52) | 67% (59, 73) | 70% (62, 75) |

[b] Total number of doses administered before study inclusion regardless timing with respect to prior primary infection (before and/or after primary infection)

[a] Logistic regression models adjusted for age, sex, type of employment, facility, indication for testing and epidemiological week. Exposures are compared to unvaccinated individuals without infection history.

Abbreviations: CI, confidence interval; PI, primary infection

**Supplementary Table 2. Protection against Omicron BA.2 reinfection induced by primary pre-Omicron or Omicron BA.1 infection with/without vaccination, by reinfection definition: ≥30-day interval versus ≥90-day interval between positive NAAT test results**

| | **Omicron BA.2 infection ≥30-day definition of reinfection** | | | | **Omicron BA.2 infection ≥90-day definition of reinfection** | | | |
|---|---|---|---|---|---|---|---|---|
| **Infection history and vaccination status[a]** | Cases | Controls | Unadjusted effectiveness (95% CI) | Adjusted effectiveness[b] (95% CI) | Cases | Controls | Unadjusted effectiveness (95% CI) | Adjusted effectiveness[b] (95% CI) |
| **Pre-Omicron PI** | 2521 | 7360 | | | 2521 | 7360 | | |
| Unvaccinated | 109 | 265 | 36% (19, 50) | 38% (19, 53) | 109 | 265 | 36% (18, 50) | 38% (18, 53) |
| 1-dose vaccinated | 342 | 729 | 27% (15, 38) | 56% (47, 63) | 342 | 727 | 27% (14, 38) | 57% (48, 64) |
| 2-dose vaccinated | 897 | 2665 | 48% (41, 54) | 69% (64, 73) | 897 | 2665 | 48% (41, 54) | 69% (64, 74) |
| 3-dose vaccinated | 1173 | 3701 | 51% (45, 56) | 70% (66, 74) | 1173 | 3703 | 51% (44, 56) | 71% (67, 75) |
| **Omicron BA.1 PI** | 659 | 12315 | | | 470 | 9118 | | |
| Unvaccinated | 125 | 727 | 73% (67, 78) | 72% (65, 78) | 91 | 604 | 77% (70, 82) | 73% (65, 79) |
| 1-dose vaccinated | 9 | 109 | 87% (75, 94) | 89% (78, 95) | 7 | 84 | 87% (72, 94) | 87% (71, 94) |
| 2-dose vaccinated | 262 | 5322 | 92% (91, 94) | 96% (95, 96) | 207 | 4290 | 93% (91, 94) | 96% (95, 96) |
| 3-dose vaccinated | 263 | 6157 | 93% (92, 94) | 96% (95, 97) | 165 | 4140 | 94% (93, 95) | 96% (95, 97) |
| **No prior infection and unvaccinated** | 672 | 1043 | Reference | Reference | 672 | 1049 | Reference | Reference |

[a] Total number of doses administered before study inclusion regardless timing with respect to prior primary infection.
[b] Logistic regression models adjusted for age, sex, type of employment, facility, indication for testing and epidemiological week. Exposures are compared to unvaccinated individuals without infection history.

Abbreviations: CI, confidence interval; NAAT, nucleic acid amplification test

**Supplementary Figure 1. Study population**

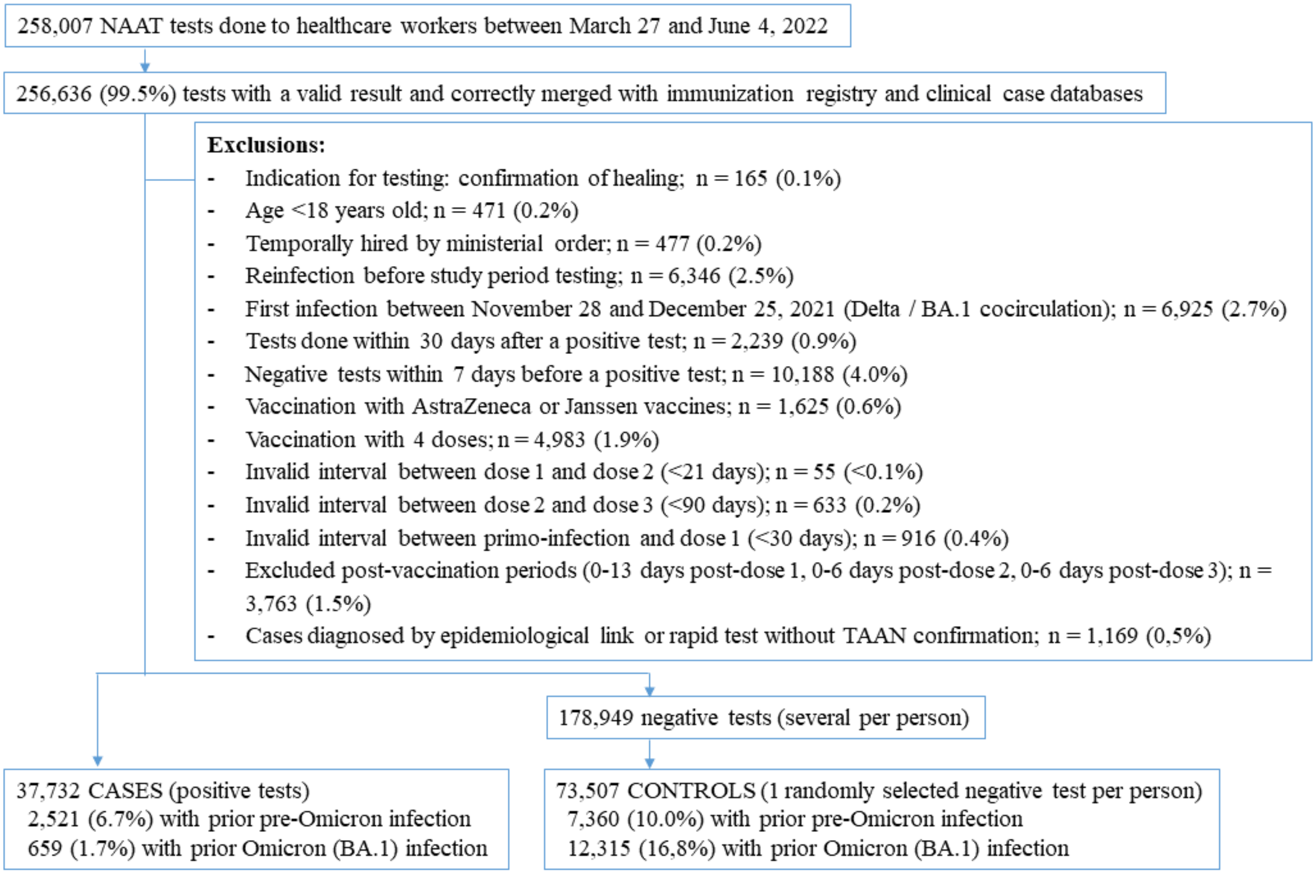

Abbreviations: NAAT, nucleic acid amplification test
